# Post-COVID-19 syndrome and related dysautonomia reduce quality of life, and increase anxiety and depressive symptoms: evidence from Greece

**DOI:** 10.1101/2023.03.05.23286811

**Authors:** Petros Galanis, Aglaia Katsiroumpa, Irene Vraka, Katerina Kosiara, Olga Siskou, Olympia Konstantakopoulou, Theodoros Katsoulas, Parisis Gallos, Daphne Kaitelidou

**Affiliations:** Clinical Epidemiology Laboratory, Faculty of Nursing, National and Kapodistrian University of Athens, Athens, Greece; Department of Radiology, P. & A. Kyriakou Children’s Hospital, Athens, Greece; Department of Tourism Studies, University of Piraeus, Piraeus, Greece; Center for Health Services Management and Evaluation, Faculty of Nursing, National and Kapodistrian University of Athens, Athens, Greece; Faculty of Nursing, National and Kapodistrian University of Athens, Athens, Greece

**Keywords:** post-COVID-19 syndrome, dysautonomia, anxiety, depression, quality of life

## Abstract

**Background:** Post-COVID-19 syndrome affects a significant number of SARS-CoV-2 infected individuals even asymptomatic cases causing several neurological and neuropsychiatric symptoms and signs.

**Materials and Methods:** An online cross-sectional study with a convenience sample was conducted in Greece from November 2022 to January 2023. We measured demographic and clinical characteristics of patients, post-COVID-19 dysautonomia, quality of life with the EQ-5D-3L, and anxiety and depressive symptoms with the Patient Health Questionnaire-4.

**Results:** Study population included 122 patients with post-COVID-19 syndrome. One out of four patients (27.8%) manifested post-COVID-19 dysautonomia, while mean duration of COVID-19 symptoms was 11.6 months. Anxiety and depressive symptoms were worse after the post-COVID-19 syndrome (p<0.001 in both cases). A statistically significant reduction in quality of life was observed among patients after the post-COVID-19 syndrome (p<0.001 for both EQ-5D-3L index value and EQ-5D-3L VAS). Post-COVID-19 dysautonomia increased depression symptoms after the post-COVID-19 syndrome (p=0.02). We found a negative relationship between duration of COVID-19 symptoms and quality of life (p<0.001). Moreover, our results showed that depressive symptoms were more often among females after the post-COVID-19 syndrome (p=0.01). Also, quality of life was lower among females than males (p=0.004 for EQ-5D-3L index value, and p=0.007 for EQ-5D-3L VAS).

**Conclusions:** Our results suggest that post-COVID-19 syndrome causes a tremendous impact on patients’ quality of life and mental health. In addition, we found that the groups most psychologically affected were patients with post-COVID-19 dysautonomia, females, and patients with longer duration of symptoms. Policy makers should attach priority to vulnerable groups in future psychiatric planning. Policy measures should focus on mental health of post-COVID-19 patients who seem to be particularly vulnerable.

## Introduction

Post-COVID-19 syndrome is defined as the condition where patients report symptoms and signs 12 or more weeks following SARS-CoV-2 infection that cannot be explained by an alternative diagnosis [1]. More than 30% of COVID-19 patients [2] and almost 80% of hospitalized COVID-19 patients [3] may experience a wide range of post-COVID-19 sequelae. According to meta-analyses, the prevalence of fatigue among patients with post-COVID-19 syndrome was 37%, the prevalence of brain fog was 32%, the prevalence of memory issues was 27%, and the prevalence of cognitive impairment was 22% [4,5]. Moreover, there are several other neurological and neuropsychiatric post-COVID-19 symptoms, such as attention disorder, anosmia, dysgeusia, headache, sleep disturbances, etc. Post-COVID-19 patients report an average of 14 persistent symptoms six months after the infection [6]. Therefore, post-COVID-19 syndrome is a heterogeneous condition with persistent symptoms and signs affecting multiple organ systems [7,8].

Dysautonomia is defined as a malfunction of the autonomic nervous system [9] and occurs in 2.5% of patients with post-COVID-19 syndrome [10]. Orthostatic hypotension, heart rate variability, and fatigue can manifest from dysautonomia in patients with post-COVID-19 syndrome [11]. However, the most frequent cardiovascular dysautonomia is postural orthostatic tachycardia syndrome especially among young adults [12]. Numerous reports suggest the development of postural orthostatic tachycardia syndrome as part of post-COVID-19 syndrome but the aetiology remains unknown [13–16]. Dysautonomia may explain the persistent clinical symptoms and signs observed in patients with post-COVID-19 syndrome, such as postural orthostatic tachycardia syndrome and fatigue [11].

Not just the physical health, but the mental health of patients with post-COVID-19 syndrome becomes paramount. Several meta-analyses found that mental health issues are prevalent among post-COVID-19 patients [17,18]. Prevalence of depression, anxiety, and post-traumatic stress disorder in post-COVID-19 community patients was estimated as 17.3%, 17.2%, and 20.6% respectively. Situation is worse for hospitalized COVID-19 patients since the prevalence of anxiety and depression was 27.5% and 23.3% respectively [17]. We should notice the wide range in prevalence of mental health issues among studies, e.g. anxiety ranged from 6.5% to 63%, depression ranged from 4% to 31%, and post-traumatic stress disorder ranged from 12.1% to 46.9% [18].

In addition, myalgia, headache, and sleep disturbances are common among patients with post-COVID-19 syndrome reducing quality of life [5]. A recent meta-analysis found that 59% of post-COVID-19 patients experienced a poor quality of life [19]. Moreover, among post-COVID-19 patients, 36%, 28%, and 8% reported extreme problems on mobility, usual activities, and self-care respectively.

To the best of our knowledge, the available studies have investigated quality of life, anxiety and depressive symptoms only after the post-COVID-19 patients. Also, there is no study that has investigated the impact of post-COVID-19 dysautonomia on quality of life, anxiety and depression among patients. Therefore, we measured quality of life, anxiety and depressive symptoms among patients before and after the post-COVID-19 syndrome in order to identify differences. Moreover, we assessed the impact of post-COVID-19 dysautonomia, demographic, and clinical characteristics on patients’ life.

## Materials and Methods

### Study design

A cross-sectional study with a convenience sample was conducted in Greece. Participants were obtained from the Long COVID Greece patients’ society [20]. This society is a non-profit organization and it is a member of a European network of Long COVID patient association (i.e., Long COVID Europe) [21].

First, we used Google forms to create an online version of the study questionnaire. The administrators of the Facebook page of the Long COVID Greece patients’ society gave us their permission to post online the study questionnaire. Thus, patients who belong to the Long COVID Greece patients’ society could participate in our study. We collected data from November 2022 to January 2023.

In order to assess changes in anxiety, depression, and quality of life among patients with post-COVID-19 syndrome we asked participants to consider their lives in two different times; before the post-COVID-19 syndrome and the time they completed the questionnaire. In that way, we measured anxiety, depression, and quality of life among patients before and after the post-COVID-19 syndrome and made comparisons.

We applied the following inclusion criteria: adults patients over 18 years old; SARS-CoV-2 infection with a confirmed molecular test; patients who met the definition of post-COVID-19 syndrome, i.e. symptoms and signs consistent with COVID-19 for more than 12 weeks after the diagnosis and cannot be attributable to alternative diagnoses [1,22].

### Measurements

First, we measured demographic and clinical characteristics of patients, i.e. gender, age, hospitalization in COVID-19 ward and COVID-19 intensive care unit, duration of COVID-19 symptoms (in months), post-COVID-19 dysautonomia, and anxiety disorders and depression before the post-COVID-19 syndrome.

We measured quality of life with the 3-level version of EQ-5D (EQ-5D-3L) [23]. This tool includes five dimensions, i.e. mobility, self-care, usual activities, pain/discomfort and anxiety/depression. Answers on these dimensions can be “no problems”, “some problems”, and “extreme problems”. All answers are converted into a single summary index value with higher values indicating better quality of life. In our study, we used the Greek set of weights in order to calculate the EQ-5D-3L index value [24]. Also, we used the EQ visual analogue scale (EQ VAS) to measure patients’ quality of life. EQ VAS takes values from 0 (worst quality of life) to 100 (best quality of life). Reliability of the EQ-5D-3L in our study was good since Cronbach’s coefficient alpha was 0.791.

We measured patients’ anxiety and depressive symptoms with the Patient Health Questionnaire-4 (PHQ-4) [25]. PHQ-4 measures anxiety with two items, and depression with two other items. Answers are on four-point Likert scale: not at all (0), a few days (1), most of the days (2), almost every day (3). A total score from 0 to 6 is calculated for anxiety and depression with higher values indicates higher levels of anxiety and depression symptoms. Individuals with anxiety or depression score ≥ 3 are probable to experience major anxiety or depression disorder. We used the Greek version of the PHQ-4 [26]. In our study, reliability of the PHQ-4 was very good since Cronbach’s coefficient alpha for the anxiety was 0.854, and the depression was 0.849.

### Ethical issues

We applied the guidelines of the Declaration of Helsinki in our study. Also, we obtained the approval by the Ethics Committee of Faculty of Nursing, National and Kapodistrian University of Athens (reference number; 420, 10 October 2022). Moreover, data collection was performed in an anonymous and voluntary basis since we did not collect personal information data. Participants gave their informed consent before they completed the study questionnaire.

### Statistical analysis

We use numbers and percentages to present categorical variables, and mean, standard deviation, median, and range to present continuous variables. Kolmogorov-Smirnov test and Q-Q plots showed that continuous variables followed normal distribution. We used repeated measures analysis of variance in order to assess differences regarding anxiety, depression, and quality of life among patients before and after the post-COVID-19 syndrome. Also, we measured the impact of dysautonomia, demographic and clinical variables on anxiety, depression, and quality of life using repeated measures analysis of variance. We used McNemar’s test to assess differences regarding possible anxiety and depression disorder, and five dimensions of the EuroQol-5D-3L before and after the post-COVID-19 syndrome. We did not assess the impact of hospitalization in COVID-19 intensive care unit in patients’ life since only four patients have been hospitalized. P-values less than 0.05 were considered as statistically significant. Statistical analysis was performed with the IBM SPSS 21.0 (IBM Corp. Released 2012. IBM SPSS Statistics for Windows, Version 21.0. Armonk, NY: IBM Corp.).

## Results

### Demographic and clinical characteristics

Study population included 89 females (73%) and 33 males (27%). Mean age was 44.8 years (standard deviation=11.5) with a range from 21 to 88 years (median=43).

Clinical characteristics of patients with post-COVID-19 syndrome are presented in Table 1. Among participants, 17.2% have been hospitalized in COVID-19 ward and 3.3% in COVID-19 intensive care unit. Mean duration of COVID-19 symptoms was 11.6 months (standard deviation=8.6) with a range from 1 to 26 months (median=8). One out of four patients (27.8%) manifested post-COVID-19 dysautonomia. In our sample, 18.9% suffered from anxiety disorders before post-COVID-19 syndrome and 11.5% have been diagnosed with depression.

**Table 1.** Clinical characteristics of patients with post-COVID-19 syndrome.

| **Variables** | **N** | **%** |
| --- | --- | --- |
| Hospitalization in COVID-19 ward |  |  |
| No | 101 | 82.8 |
| Yes | 21 | 17.2 |
| Hospitalization in COVID-19 intensive care unit |  |  |
| No | 118 | 96.7 |
| Yes | 4 | 3.3 |
| Duration of COVID-19 symptoms (months), mean, standard deviation | 11.6 | 8.6 |
| Post-COVID-19 dysautonomia |  |  |
| No | 65 | 72.2 |
| Yes | 25 | 27.8 |
| Anxiety disorders before post-COVID-19 syndrome |  |  |
| No | 99 | 81.1 |
| Yes | 23 | 18.9 |
| Depression before post-COVID-19 syndrome |  |  |
| No | 108 | 88.5 |
| Yes | 14 | 11.5 |

### Anxiety, depression, and quality of life

Anxiety, depression, and quality of life before and after the post-COVID-19 syndrome are presented in Table 2. Anxiety and depressive symptoms were worse after the post-COVID-19 syndrome (p<0.001 in both cases). In particular, mean anxiety score before the post-COVID-19 syndrome was 1.41 and after the syndrome was 3.22. In a similar way, mean depression score before the post-COVID-19 syndrome was 0.81 and after the syndrome was 3.56.

**Table 2.** Anxiety, depression, and quality of life before and after the post-COVID-19 syndrome.

| Scale | Post-COVID-19 syndrome |  | P-value<sup>a</sup> |
| --- | --- | --- | --- |
|  | Before | After |  |
| Patient Health Questionnaire |  |  |  |
| Anxiety score | 1.41 (1.28) | 3.22 (1.78) | <0.001 |
| Depression score | 0.81 (1.19) | 3.56 (1.70) | <0.001 |
| EQ-5D-3L index value | 0.85 (0.19) | 0.36 (0.37) | <0.001 |
| EQ-5D-3L VAS | 85.64 (13.63) | 54.10 (21.71) | <0.001 |

Values are expressed as mean (standard deviation)
<sup>a</sup> repeated measures analysis of variance

Additionally, only 12.3% (n=15) of patients had anxiety score ≥ 3 before the post-COVID-19 syndrome, while the respective percentage after the syndrome was 60.7% (n=74) (p<0.001) indicating possible major anxiety disorder. Similarly, 69.7% (n=85) of patients had depression score ≥ 3 after the post-COVID-19 syndrome, while before the syndrome the respective percentage was only 9% (n=11) indicating possible major depression disorder.

A statistically significant reduction in quality of life was observed among patients after the post-COVID-19 syndrome (p<0.001 for both EQ-5D-3L index value and EQ-5D-3L VAS), (Table 2). EQ-5D-3L index value reduced by 57.6% (0.85 vs. 0.36), while EQ-5D-3L VAS reduced by 36.8% (85.64 vs. 54.10). Descriptive statistics for the five dimensions of the EQ-5D-3L before and after the post-COVID-19 syndrome are shown in Table 3. Prevalence of problems was higher in all dimensions of the EQ-5D-3L (p<0.001 in all cases). Specifically, 9% and 79.5% of patients had mobility problems before and after the post-COVID-19 syndrome respectively. Prevalence of self-care problems was 2.5% before the post-COVID-19 syndrome and 37.7% after the syndrome. Before the post-COVID-19 syndrome only 4.9% of patients have problems with performing usual activities but this percentage was 80.3% after the syndrome. Prevalence of pain/discomfort was more than twice as high after the post-COVID-19 syndrome (79.5% vs. 30.3%). Before the post-COVID-19 syndrome 42.6% of patients felt anxious or depressed, while after the syndrome 86.1% felt like this.

**Table 3.** Descriptive statistics for the five dimensions of the EuroQol-5D-3L before and after the post-COVID-19 syndrome.

| Dimensions | Post-COVID-19 syndrome |  |  |  | P-value<sup>a</sup> |
| --- | --- | --- | --- | --- | --- |
|  | Before |  | After |  |  |
|  | N | % | N | % |  |
| Mobility |  |  |  |  | <0.001 |
| No problems | 111 | 91.0 | 25 | 20.5 |  |
| Any problems | 11 | 9.0 | 97 | 79.5 |  |
| Self-care |  |  |  |  | <0.001 |
| No problems | 119 | 97.5 | 76 | 62.3 |  |
| Any problems | 3 | 2.5 | 46 | 37.7 |  |
| Usual activities |  |  |  |  | <0.001 |
| No problems | 116 | 95.1 | 24 | 19.7 |  |
| Any problems | 6 | 4.9 | 98 | 80.3 |  |
| Pain/discomfort |  |  |  |  | <0.001 |
| No problems | 85 | 69.7 | 25 | 20.5 |  |
| Any problems | 37 | 30.3 | 97 | 79.5 |  |
| Anxiety/depression |  |  |  |  | <0.001 |
| No problems | 70 | 57.4 | 17 | 13.9 |  |
| Any problems | 52 | 42.6 | 105 | 86.1 |  |

<sup>a</sup> McNemar's test

### Relationships with dysautonomia, demographic and clinical characteristics

We present relationships between post-COVID-19 dysautonomia, demographic and clinical characteristics and anxiety, depression, and quality of life in Table 4.

**Table 4.** Anxiety, depression, and quality of life before and after the post-COVID-19 syndrome according to demographic and clinical variables of patients.

| Variables | Anxiety score |  |  | Depression score |  |  | EQ-5D-3L index value |  |  | EQ-5D-3L VAS |  |  |
| --- | --- | --- | --- | --- | --- | --- | --- | --- | --- | --- | --- | --- |
|  | Before | After | P-value<sup>a</sup> | Before | After | P-value<sup>a</sup> | Before | After | P-value<sup>a</sup> | Before | After | P-value<sup>a</sup> |
| Gender |  |  | 0.13 |  |  | 0.01 |  |  | 0.004 |  |  | 0.007 |
| Females | 1.43 (1.20) | 3.38 (1.76) |  | 0.76 (1.07) | 3.78 (1.72) |  | 0.85 (0.17) | 0.30 (0.36) |  | 86.22 (12.82) | 51.41 (21.82) |  |
| Males | 1.36 (1.48) | 2.78 (1.80) |  | 0.94 (1.50) | 2.97 (1.51) |  | 0.83 (0.24) | 0.49 (0.34) |  | 84.09 (15.71) | 61.37 (19.94) |  |
| Age (years) |  |  | 0.99 |  |  | 0.46 |  |  | 0.80 |  |  | 0.23 |
| ≤43 | 1.73 (1.51) | 3.54 (1.73) |  | 1.08 (1.41) | 3.70 (1.67) |  | 0.83 (0.21) | 0.35 (0.34) |  | 87.68 (12.59) | 53.78 (22.26) |  |
| >43 | 1.07 (0.85) | 2.88 (1.79) |  | 0.53 (0.84) | 3.41 (1.72) |  | 0.86 (0.16) | 0.36 (0.39) |  | 83.48 (14.46) | 54.44 (21.29) |  |
| Hospitalization in COVID-19 ward |  |  | 0.41 |  |  | 0.31 |  |  | 0.52 |  |  | 0.20 |
| No | 1.31 (1.25) | 3.06 (1.81) |  | 0.77 (1.20) | 3.44 (1.72) |  | 0.87 (0.18) | 0.37 (0.38) |  | 86.73 (13.23) | 54.00 (22.77) |  |
| Yes | 1.91 (1.30) | 4.00 (1.48) |  | 1.00 (1.18) | 4.14 (1.49) |  | 0.75 (0.21) | 0.31 (0.30) |  | 80.43 (14.63) | 54.62 (16.08) |  |
| Duration of COVID-19 symptoms (months) |  |  | 0.54 |  |  | 0.21 |  |  | <0.001 |  |  | <0.001 |
| ≤8 | 1.19 (1.17) | 2.91 (1.81) |  | 0.67 (1.07) | 3.20 (1.70) |  | 0.85 (0.17) | 0.47 (0.36) |  | 84.45 (14.23) | 60.17 (21.13) |  |
| >8 | 1.66 (1.36) | 3.57 (1.71) |  | 0.97 (1.31) | 3.95 (1.62) |  | 0.84 (0.21) | 0.23 (0.33) |  | 86.96 (12.94) | 47.40 (20.48) |  |
| Post-COVID-19 dysautonomia |  |  | 0.49 |  |  | 0.02 |  |  | 0.04 |  |  | 0.10 |
| No | 1.23 (1.14) | 2.75 (1.69) |  | 0.88 (1.26) | 2.94 (1.50) |  | 0.86 (0.17) | 0.46 (0.37) |  | 85.64 (13.52) | 59.51 (23.73) |  |
| Yes | 1.64 (1.41) | 3.44 (2.12) |  | 0.56 (1.20) | 3.64 (1.91) |  | 0.81 (0.23) | 0.26 (0.35) |  | 85.00 (15.28) | 50.20 (16.04) |  |
| Anxiety disorders before post-COVID-19 syndrome |  |  | 0.72 |  |  | 0.57 |  |  | 0.92 |  |  | 0.76 |
| No | 1.16 (1.09) | 3.00 (1.73) |  | 0.73 (1.22) | 3.43 (1.70) |  | 0.89 (0.15) | 0.41 (0.35) |  | 75.35 (19.67) | 56.79 (21.56) |  |
| Yes | 2.48 (1.50) | 4.17 (1.75) |  | 1.13 (1.06) | 4.09 (1.62) |  | 0.65 (0.22) | 0.15 (0.37) |  | 75.35 (19.67) | 42.52 (18.68) |  |
| Depression before post-COVID-19 syndrome |  |  | 0.38 |  |  | 0.62 |  |  | 0.44 |  |  | 0.77 |
| No | 1.27 (1.21) | 3.13 (1.76) |  | 0.74 (1.16) | 3.52 (1.71) |  | 0.87 (0.17) | 0.39 (0.34) |  | 87.35 (12.22) | 56.02 (21.07) |  |
| Yes | 2.50 (1.29) | 3.93 (1.90) |  | 1.36 (1.39) | 3.86 (1.66) |  | 0.64 (0.22) | 0.08 (0.43) |  | 72.50 (17.07) | 39.29 (21.56) |  |

Values are expressed as mean (standard deviation)
<sup>a</sup> repeated measures analysis of variance

Post-COVID-19 dysautonomia increased depression symptoms after the post-COVID-19 syndrome (p=0.02). In particular, depression score increased by 2.08 units among patients without dysautonomia, and by 3.08 units among patients with dysautonomia. Moreover, post-COVID-19 dysautonomia decreased quality of life significantly (p=0.04). In that case, EQ-5D-3L index value reduced by 50% (0.86 vs. 0.37) among patients without dysautonomia, and by 67.9% (0.81 vs. 0.26) among patients with dysautonomia.

Among other variables, we found a negative relationship between duration of COVID-19 symptoms and quality of life (p<0.001 for both EQ-5D-3L index value and EQ-5D-3L VAS). Among patients who had COVID-19 symptoms less than eight months, EQ-5D-3L index value decreased from 0.85 to 0.47 (0.38 units), while among patients who had COVID-19 symptoms more than eight months we found a decrease from 0.84 to 0.23 (0.61 units).

Also, we found that females experienced more difficulties after the post-COVID-19 syndrome. In particular, depression score increased by 3.02 units among females but by 2.03 among males after the post-COVID-19 syndrome (p=0.01). Moreover, EQ-5D-3L index value decreased from 0.85 to 0.30 (0.55 units) among females, and from 0.83 to 0.49 (0.34 units) among males (p=0.004). A similar trend was observed for EQ-5D-3L VAS (p=0.007).

## Discussion

We performed a study to assess the impact of post-COVID-19 syndrome and related dysautonomia on patients’ life. To the best of our knowledge, our study measured for first time quality of life, anxiety and depressive symptoms among patients before and after the post-COVID-19 syndrome. Additionally, we were the first that investigated the impact of post-COVID-19 dysautonomia on patients’ quality of life, anxiety and depression.

We found a tremendous decrease in quality of life in post-COVID-19 patients. In particular, EQ-5D-3L index value among our patients removed from 0.85 before the post-COVID-19 syndrome to 0.36 after the syndrome. Since mean age of our sample was 44.8 years we compared our results with the 45-54 group and we found that the Greek EQ-5D-3L index norm value is 0.916 [27], while in our study was only 0.36 after the post-COVID-19 syndrome. Also, the Greek EQ-5D-3L index norm value in the ≥ 75 group is twice as high the EQ-5D-3L index value in our patients (0.74 vs. 0.36). A similar trend was found in case of EQ-5D-3L VAS. In particular, EQ-5D-3L VAS decreased from 85.64 to 54.10 after the post-COVID-19 syndrome, while the Greek EQ-5D-3L VAS norm value is 78 in the 45-54 group and 54 in the ≥ 75 group. Additionally, data from other countries (i.e. France, Spain, USA, and China) suggest that the EQ-5D-3L VAS in post-COVID-19 patients ranged from 64 to 80, while the EQ-5D-3L index value ranged from 0.71 to 0.86 [28–31]. Moreover, our results showed that the majority of patients experienced mobility problems (79.5%), usual activities problems (80.3%), pain/discomfort (79.5%), and anxiety/depression (86.1%) according to the EQ-5D-3L. A meta-analysis including studies until March 2021 which used the EQ-5D-3L found that post-COVID-19 patients experienced less mobility problems (36%), usual activities problems (28%), pain/discomfort (42%), and anxiety/depression (38%) than our patients [19]. These big differences could be attributed to the fact that early studies on post-COVID-19 syndrome included mainly patients who experience the first months of the syndrome, while our patients suffered from the syndrome for a prolonged time since the mean duration of COVID-19 symptoms was about a year.

We observed a similar trend with quality of life regarding anxiety and depression in our sample. In particular, patients reported a huge difference in anxiety and depressive symptoms before and after the post-COVID-19 syndrome. Our results indicated that 60.7% of post-COVID-19 patients may suffer from a major anxiety disorder, and 69.7% may suffer from a major depression disorder. A meta-analysis including studies until July 2021 found that the prevalence of anxiety (23%) and depression (16.7%) among post-COVID-19 patients were much lower than our study [5]. As we notice above, it is probable a relation between the prevalence of mental health issues and the duration of post-COVID-19 syndrome. Early studies support this hypothesis since they found that the frequency of anxiety, depression, fatigue, brain fog, and insomnia increased from mid- to long-term follow up [32,33]. Moreover, according to a meta-analysis, post-COVID-19 patients with worse clinical course (e.g. hospitalized patients) experienced higher prevalence of anxiety, depression, fatigue, and sleep disturbances [5]. These findings are in line with our results since we found a negative relationship between duration of COVID-19 symptoms and quality of life.

As we hypothesized, post-COVID-19 dysautonomia reduced quality of life in our patients and increased depressive symptoms. Since our study was the first that attempt to estimate the relationship between post-COVID-19 dysautonomia and patients’ life we cannot make direct comparisons with the literature. However, several studies found that patients with post-COVID-19 dysautonomia experienced higher levels of fatigue [6,11,19,34]. Moreover, post-COVID-19 dysautonomia was associated with objective functional limitations, such as reduced work rate and exercise [35]. Thus, work capacity and energy of patients with dysautonomia seem to be restricted. These findings are unsurprising since fatigue is described as a major clinical feature of post-COVID-19 dysautonomia [34]. Therefore, it is probable that high levels of fatigue among post-COVID-19 patients limit usual activities of patients, such as personal and work life, and family responsibilities. Then, post-COVID-19 patients may feel less active, lose their passion and motivation, and avoid their family members and friends resulting on an increase of depressive symptoms and a decline in quality of life.

Among demographic and clinical variables we found that gender affected depression and quality of life. In particular, our results showed that depressive symptoms were more often among females after the post-COVID-19 syndrome. Also, quality of life was lower among females than males. This finding is confirmed by the literature since several reviews found that female patients had a poorer quality of life, greater disability, and higher prevalence of depression and anxiety [36–38]. Moreover, during the pandemic COVID-19 patients were more likely to be depressed, and anxious, while quality of life was worse for females [39–42].

Our study had several limitations. First, we did not measure participants’ quality of life, anxiety, and depression before the post-COVID-19 syndrome since it was impossible to predict post-COVID-19 patients. Instead, we asked post-COVID-19 patients to rate their quality of life, anxiety, and depression before the occurrence of post-COVID-19 syndrome. Thus, a recall bias was probable in our study. Second, our cross-sectional study can assess patients’ life in a particular moment. Therefore, several conditions such as the relapse of syndrome or the rebound effect could affect our results introducing information bias. Longitudinal studies measuring quality of life, anxiety, and depression in different times could add invaluable information. Third, we used self-reported questionnaires to measure our variables and an information bias was probable. Fourth, we used a convenience and small sample since it is difficult to approach post-COVID-19 patients who experience a new condition where information and knowledge is limited even among healthcare workers. Further studies with more representative and bigger samples should be conducted. Finally, we measured the impact of several demographic and clinical variables but there are many other factors that could have an impact on post-COVID-19 patients’ life and should be investigated in the future.

## Conclusions

Our results suggest that post-COVID-19 syndrome causes a tremendous impact on patients’ quality of life and mental health. In addition, we found that the groups most psychologically affected by the post-COVID-19 syndrome were patients with post-COVID-19 dysautonomia, females, and patients with longer duration of symptoms. Therefore, we should prioritize our efforts to help post-COVID-19 patients since they fight an unknown situation without effective therapeutic interventions. Research on post-COVID-19 syndrome is evolving but our knowledge until now is limited. Further research on the psychological impact of post-COVID-19 syndrome is recommended especially among vulnerable groups such as patients with post-COVID-19 dysautonomia and females in order to get more valid results. Policy makers should attach priority to vulnerable groups in future psychiatric planning. Policy measures should focus on mental health of post-COVID-19 patients who seem to be particularly vulnerable. Healthcare workers should improve their knowledge on post-COVID-19 syndrome taking into their consideration that mental health of post-COVID-19 patients is compromised considerably.

## Data Availability

All data produced in the present study are available upon reasonable request to the authors

## Acknowledgments

The authors would like to thank the clinicians and Long COVID Greece society members who contributed their time, thoughts, and experiences to this study.

